# Revisiting the Links Between Asthma and the Psychosis Spectrum: shared molecular mechanisms

**DOI:** 10.1101/2025.08.27.25334529

**Authors:** Christina Dardani, Jamie W. Robinson, Alexandra Havdahl, Liza Darrous, Hannah J. Jones, Stan Zammit, Sarah A. Sullivan, Dheeraj Rai, Evie Stergiakouli, Zhaozhong Zhu, Liming Liang, George W. Nava, Renee Gardner, Jakob Grove, Tom G. Richardson, George Davey Smith, James W. Dodd, Gibran Hemani, Tom R. Gaunt, Golam M. Khandaker

## Abstract

Epidemiological studies suggest associations of asthma with the psychosis spectrum (psychotic experiences, bipolar disorder, schizophrenia), but the mechanisms underlying these associations remain unclear. We examined the relationship between asthma and psychosis-related outcomes using observational cohort, polygenic score, and Mendelian randomization (MR) analyses, and assessed the possibility of shared genetic underpinnings between these traits using genetic colocalisation. Results from a UK population-based prospective birth cohort suggest that asthma at age 7 and polygenic risk for asthma are associated with psychotic experiences in early adulthood. Results from two-sample MR analyses do not support causal relationships of genetic liability to asthma with bipolar disorder or schizophrenia. Instead, genetic correlation and colocalization analyses point to the presence of shared genetic aetiology between the conditions. We identified 8 genomic regions with potentially shared causal genes between asthma and bipolar disorder or asthma and schizophrenia. Using genetically predicted mRNA expression in condition-relevant tissues (brain and lung), we identified 16 genes shared between conditions, which include *FADS1*, *SLC4A10, BDH2,* and *CISD2*. Our results suggest that population-level associations of asthma with psychosis spectrum conditions could be due to shared molecular mechanisms involving fatty acid metabolism, ion channel activity, and iron homeostasis.

## Background

Asthma affects approximately 300 million people worldwide^1^. It is characterised by chronic inflammation of the respiratory airways causing recurrent episodes of wheeze, shortness of breath, chest tightness and cough^2^. Emerging evidence suggests that individuals with asthma are at increased risk of adverse life outcomes^3^, including chronic mental health conditions^4^. Among notable mental health comorbidities, existing research suggests strong links of asthma with psychotic disorders, a heterogeneous group of serious neuropsychiatric conditions that are among the leading causes of disability worldwide^5^.

Psychotic disorders, or psychosis, is an umbrella term encompassing a range of conditions that affect approximately 3% of the population and involve mainly atypical perception and cognition^6^. Psychotic disorders may or may not have a mood element, thus these are often grouped as non-affective (e.g., schizophrenia – the archetypal psychotic disorder) and affective (e.g., bipolar disorder). Psychotic symptoms or experiences (e.g., hallucinations) are reported by about 5% of adults in the population and are considered part of the psychosis spectrum (Figure 1)^7^. Largescale population studies suggest associations of asthma with conditions and traits across the psychosis spectrum. Longitudinal nationwide cohort studies from Taiwan, Denmark, and Sweden have reported associations of asthma in childhood/adolescence with bipolar disorder^8,9^ and schizophrenia^9–11^ in adulthood. Furthermore, associations between asthma and subsequent psychotic experiences have been reported from studies involving retrospective recall of past asthma (cross-sectional data from 16 countries)^12^ and prospective assessment of childhood asthma (birth cohort studies)^13^.

**Figure 1.**
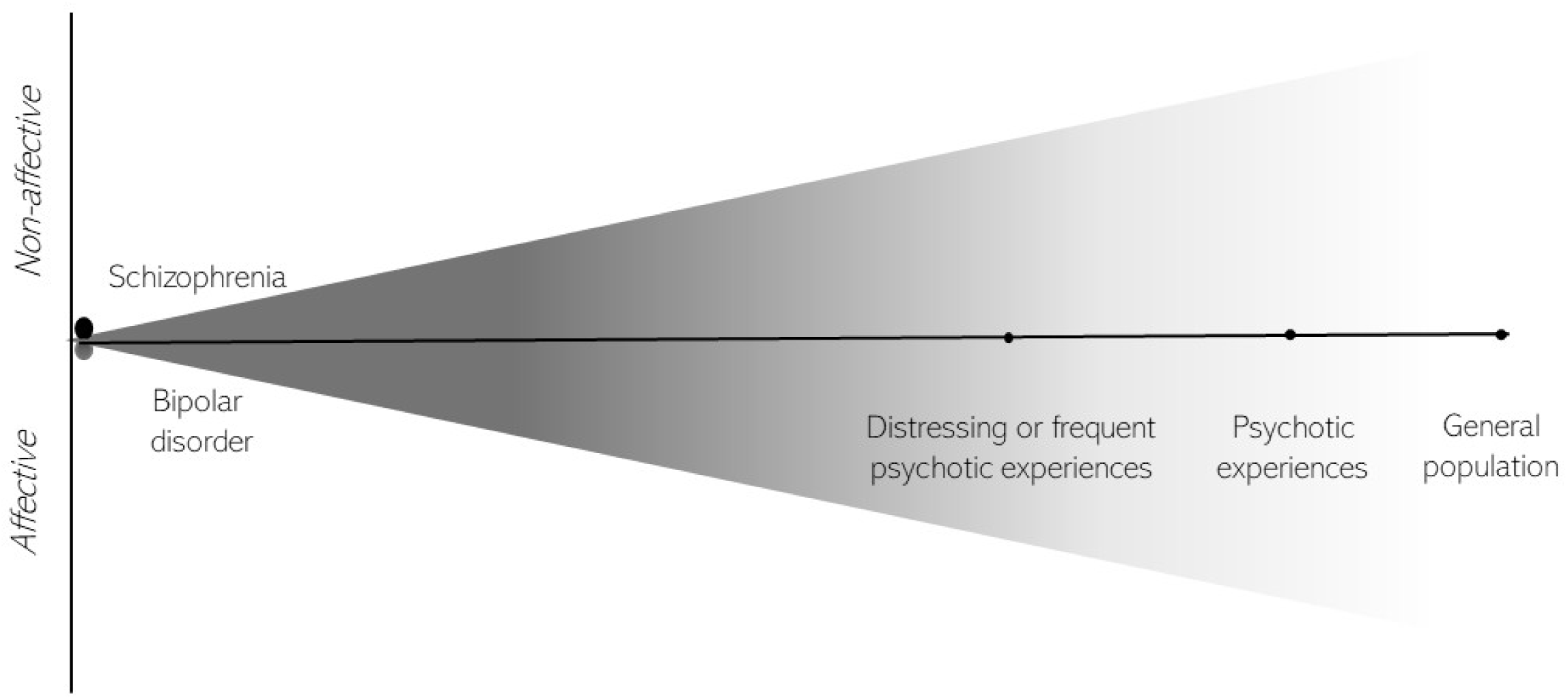
Visual representation of the psychosis spectrum in which schizophrenia (non-affective psychosis) and bipolar disorder (affective psychosis) lie at the clinically severe end (left) and psychotic experiences span to the general population (right). The figure is for illustrative purposes only and does not reflect all conditions residing in the spectrum.

The reasons underlying previously reported links of asthma to the psychosis spectrum are likely complex, with environmental and genetic factors contributing to the co-occurrence. For example, corticosteroids (among the first-line treatments for asthma) are associated with a number of adverse psychiatric effects including psychosis and mood alterations^14,15^. However, register-based studies in Taiwan and Denmark found that the asthma-psychosis associations remain even after taking into account corticosteroid use, suggesting that some biological components of asthma itself might, at least partially, contribute to psychotic disorders risk^11,16^. Indeed, a Swedish register-based study found associations between both maternal and paternal history of asthma (pre-pregnancy) and offspring diagnosis of bipolar disorder, suggesting a potential role of underlying genetics^9^. This possibility is further supported by genetic correlations between asthma and bipolar disorder^17^. However, Mendelian randomization (MR) studies did not find direct causal effects of genetic liability to asthma on either bipolar disorder or schizophrenia. Therefore, it is possible that the comorbidity between asthma and the psychosis spectrum conditions is driven by specific shared genetic variants, but which those remain unknown.

This study has two objectives. First, we revisit and update epidemiological evidence on the links between asthma and psychosis spectrum traits using population-based and genetic data. This involves: (1.1) cohort analyses investigating the prospective associations between asthma in childhood and psychotic experiences assessed in adulthood; (1.2) polygenic risk score (PRS) analyses interrogating the associations between genetic liability to asthma and psychotic experiences assessed in adulthood; and (1.3) MR analyses assessing evidence of causality in the associations of asthma with schizophrenia and bipolar disorder. Second, we elucidate shared underlying genetic mechanisms between asthma, schizophrenia and bipolar disorder using genomic and transcriptomic data. This involves: (2.1) linkage-disequilibrium score regression (LDSC) analyses assessing the genetic correlations of asthma to schizophrenia and bipolar disorder; (2.2) genetic colocalisation analyses investigating whether asthma shares causal loci with the conditions; and (2.3) tissue-specific genetic colocalisation analyses testing whether any shared loci are implicated in the expression of specific genes in lungs and brain. A visual summary of the objectives and analyses conducted in the present study can be found in Figure 2.

**Figure 2.**
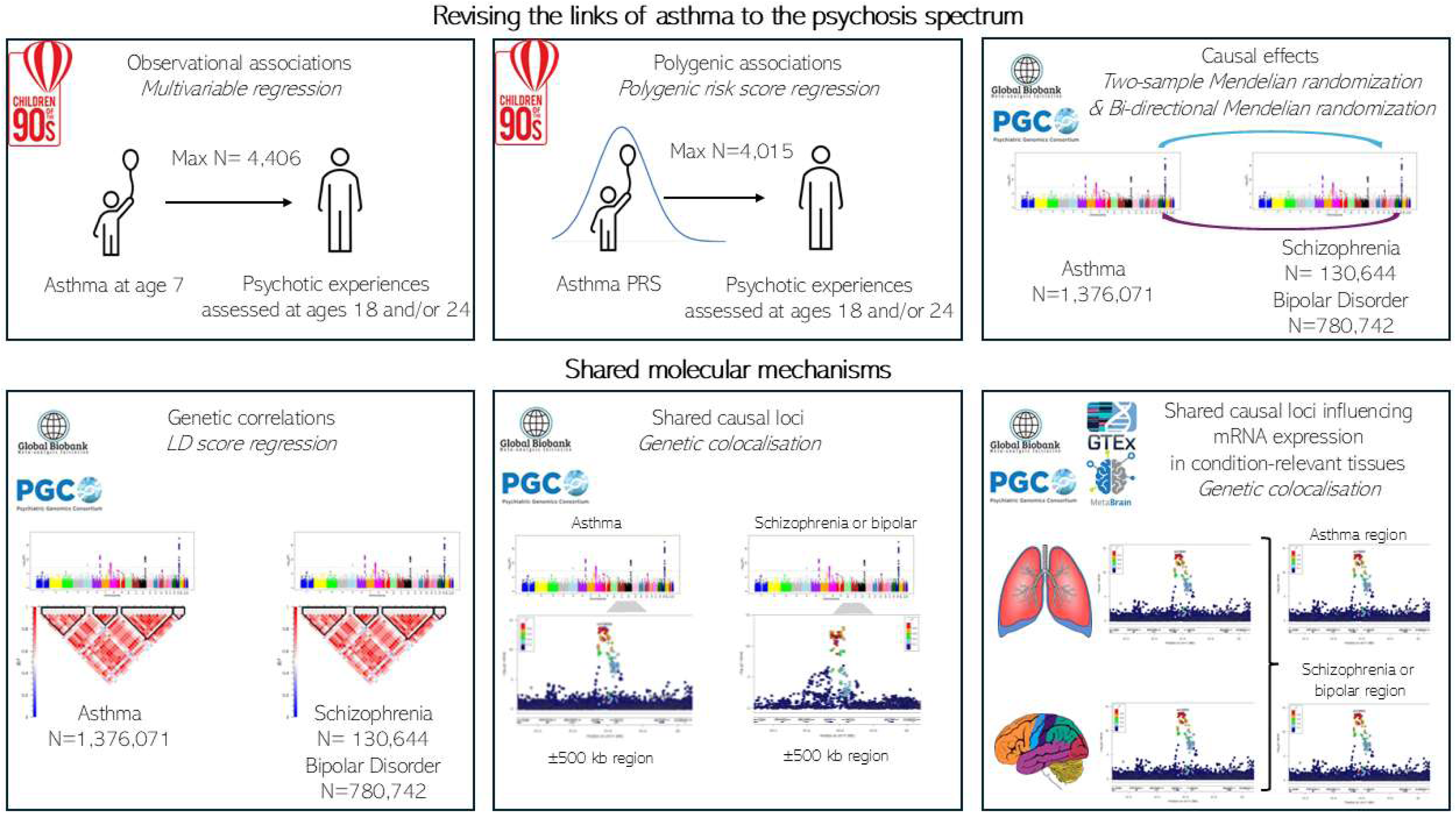
Visual summary of study objectives and analyses conducted.

## Methods

### Objective 1 Revisiting the links of asthma to the psychosis spectrum

#### (1.1) Longitudinal cohort study of childhood asthma and subsequent psychotic experiences

Using phenotype data from the Avon Longitudinal Study of Parents and Children (ALSPAC) cohort we tested the associations between asthma at age 7 and psychotic experiences assessed at ages 18 and/or 24.

##### Data

Between April 1991 and December 1992 more than 14,000 pregnant women were recruited into the ALSPAC birth cohort and these, the children arising from the pregnancy, and their partners have been followed up intensively over two decades. The total sample size for analyses using any data collected after the age of seven is 15,447 pregnancies, resulting in 15,658 foetuses. Of these 14,901 children were alive at 1 year of age. Details on the ALSPAC cohort can be found in Supplementary Note 1, on the ALSPAC website (http://www.bristol.ac.uk/alspac) and elsewhere^20–22^. The study website contains details of all the data that is available through a fully searchable data dictionary and variable search tool (http://www.bristol.ac.uk/alspac/researchers/our-data/). Ethical approval for the study was obtained from the ALSPAC Ethics and Law Committee and the Local Research Ethics Committees (B4198).

##### Exposures

Asthma: ALSPAC mothers completed a postal questionnaire indicating whether a doctor had ever diagnosed their child with asthma when the child was approximately 7.5 years old. The measure has been previously validated against general practitioner (GP) recorded asthma diagnosis and was found have good sensitivity (88.5%) and specificity (95.7%)^23^.

##### Outcomes

Psychotic experiences: Psychotic experiences were assessed at ages 18 and 24 using the semi structured Psychosis-Like Symptoms interview (PLIKSi), administered by trained psychologists, and scored according to criteria predefined by the World Health Organization^24^. The PLIKSi consists of 12 core questions covering hallucinations, delusions, and thought interference. At the age 24 assessment, questions also asked about tactile hallucinations (total 13 questions-please refer to Supplementary Note 2 for further details on the PLIKSi). Participants were asked about experiences that had occurred since age 12, and those that had occurred over the past 6 months prior to the assessment. Psychotic experiences were considered as present if they were rated by the interviewer as suspected or definitely present, and not attributable to falling asleep, waking up, or fever. Our primary outcome was psychotic experiences assessed at ages 18 and/or 24 that were reported to have occurred since age 12. Additionally, we examined psychotic experiences that were reported as distressing and/or frequent at the ages 18 and/or 24 assessment, since these experiences could be more clinically relevant and predictive of psychotic disorder^25^. In sensitivity analyses, to minimise potential recall bias, we used two further outcomes covering a shorter timeframe: psychotic experiences assessed at age 18 that were reported to have occurred over the past 6 months, and psychotic experiences assessed at age 24 that were reported to have occurred over the past 6 months.

##### Covariates

We included sex at birth (male/female), maternal parity (1 child versus ≥ 2 children), major financial problems in the family when the child was 8 months old (yes/no), maternal highest educational attainment (32 weeks gestation), smoking in the house when the child was approximately 8 years old (not smoking in the house vs smoking), maternal Crown-Crisp anxiety scores (18 weeks gestation)^26^, maternal depression measured with the Edinburgh Postnatal Depression Scale (EPDS; 18 weeks gestation; dichotomised scores ≥ 12)^27^, and child IQ scores at age 8 assessed with the Wechsler Intelligence Scale for Children third edition (WISC-III)^28^.

##### Analyses

We estimated descriptive statistics of participant characteristics for individuals with and without asthma at age 7 and psychotic experiences. Using logistic regression, we estimated odds ratios (ORs) and 95% confidence intervals (CIs) for the associations between asthma in childhood and psychotic experiences assessed in adulthood. We performed crude models and covariate-adjusted models.

#### (1.2) Polygenic associations between asthma and psychotic experiences in the ALSPAC cohort

Using genotype data from the ALSPAC cohort we tested the associations between PRSs for asthma and psychotic experiences assessed at ages 18 and/or 24.

##### Data

Discovery sample: We used summary data from the latest genome-wide association study (GWAS) of asthma comprising 1,376,071 individuals (Ncases= 121,940; Ncontrols= 1,254,131) of European ancestry from 14 cohorts (BioMe, BioVU, CCPM, DECODE, ESTBB, FinnGen, GS, HUNT, Lifelines, MGB, MGI, QSKIN, UCLA, UKBB). Please see original publication for further details^29^.

Target sample: We used individual-level genotype data from the ALSPAC cohort. A total of 9,912 ALSPAC children were genotyped on the Illumina HumanHap550 quad chip genotyping platforms by 23andme subcontracting the Wellcome Trust Sanger Institute, Cambridge, UK, and the Laboratory Corporation of America, Burlington, NC, USA. After quality control (Supplementary Note 3) and removing participants who had withdrawn consent, genotype data were available for N=7,856. Consent for biological samples were collected in accordance with the Human Tissue Act (2004).

##### Exposures

PRS for asthma: PRSs were calculated using PLINK v.1.9, applying the method described by the Psychiatric Genomics Consortium (PGC)^30^. SNPs with mismatching alleles between the discovery and target dataset were removed. The wider region of the Major Histocompatibility Complex (MHC) was removed (25–34 Mb), except for one SNP representing the strongest signal. Using ALSPAC data as the reference panel, SNPs were clumped with an r2 of 0.25 and a physical distance threshold of 500 kB. For each participant, we calculated PRS across 13 P-value thresholds (P< 5.0 × 10^−08^ to P< 0.5), standardized by subtracting the mean and dividing by the standard deviation.

##### Outcomes

Psychotic experiences: We used the four psychotic experiences outcomes described above, i.e., psychotic experiences assessed at ages 18 and/or 24, distressing and/or frequent psychotic experiences assessed at ages 18 and/or 24, psychotic experiences occurring in the past 6 months at age 18 and psychotic experiences occurring in the past 6 months at age 24.

##### Analyses

First, we performed logistic regression analyses testing the associations between each of the 13 asthma PRSs and asthma at age 7, adjusted for the child’s sex and the 10 principal components of the ALSPAC genotype data. For each model we estimated the area under receiver operating characteristic curve (AUC). The PRS with the highest AUC value was defined as our primary exposure. Second, we performed logistic regression analyses to examine the associations between the 13 PRSs for asthma and psychotic experiences outcomes up to young adulthood. Analyses were adjusted for child’s sex and the first 10 principal components of the ALSPAC genotype data. Considering evidence in the literature that suggests clumping and thresholding approaches might be prone to type 1 error, we applied the PRS-PCA approach^31^. Specifically, we performed principal component analyses across the 13 PRS thresholds and used the resulting first principal component of the PRSs to repeat the association analyses. The PRS-PCA approach has been found to minimise type 1 error bias that might influence analyses using clumping and thresholding approaches^31^.

#### (1.3) Potential causal relationships between genetic liability to asthma, schizophrenia and bipolar disorder

We performed two-sample MR analyses to assess the potential causal effects of genetic liability to asthma on schizophrenia and bipolar disorder and *vice versa* using GWAS summary data for the conditions. MR can be implemented as an instrumental variable approach, utilizing common genetic variants as instruments for exposures of interest, allowing the assessment of causal effects and their direction on outcomes^32,33^. Genetic instruments for MR analysis need to satisfy specific assumptions (Supplementary Note 4)^34^. In two-sample MR, effect sizes and standard errors of the genetic instruments for the exposure and the outcome are extracted from separate GWASs conducted in independent samples from the same underlying population^32,33^.

##### Data

We used the latest European ancestry GWAS data on asthma (described above), schizophrenia (Ncases= 53,386; Ncontrols= 77,258)^35^, and bipolar disorder (Ncases= 57,833; Ncontrols= 722,909)^36^. Please see original publications for further details^29,35,37^. For bipolar disorder, we selected the available GWAS data excluding UK Biobank to avoid sample overlap with the GWAS of asthma.

##### Genetic instruments

As genetic instruments for asthma, we selected single nucleotide polymorphisms (SNPs) that were genome-wide significant (p≤ 5 ×10^−8^) and independent (r^2^= 0.01; 10,000kb; default two-sample MR package settings). In the case of the bi-directional MR analyses (genetic liability to schizophrenia->asthma; genetic liability to bipolar disorder-> asthma) we used the same criteria to select instruments for schizophrenia and bipolar disorder.

##### Analyses

We harmonized the alleles of the outcome on the exposure, to ensure SNP-exposure and SNP-outcome effects correspond to the same allele. The primary MR analysis was the inverse variance weighted (IVW) method which provides an overall causal effect estimate of the exposure on the outcome, estimated as a meta-analysis of the ratios of the SNP-outcome effect to the SNP-exposure effect weighted by each SNP’s relative precision^38^. We assessed the consistency of the IVW effect estimates using sensitivity analyses including MR Egger regression^38^, weighted median^39^ and weighted mode^40^ (Supplementary Note 5).

As an additional sensitivity analysis, we applied an extension of MR, LHC-MR^41^, which allows the estimation of bi-directional effects between phenotypes of interest, while accounting for the potential influence of a latent heritable confounder acting on both exposure and outcome. This can be particularly relevant in the case of the asthma-schizophrenia/bipolar disorder links, considering that the phenotypes share strong genetic correlations with a number of phenotypes that might influence the identified effects (e.g., BMI^42,43^). LHC-MR is using genome-wide SNPs (rather than genome-wide significant SNPs). The method is combining structural equation modelling and MR approaches to model a latent heritable confounder and provide an estimate of its potential effect on both exposure and outcome. This is important in the context of the present study, as the latent heritable confounder represents shared underlying genetic contributions on the phenotypes of interest. In addition, the method provides a causal effect estimate between exposure and outcome while accounting for the effects of the latent heritable confounder. Further details on the method can be found in the original publication^41^.

### Objective 2 Elucidating shared molecular mechanisms between asthma and psychosis spectrum conditions

#### (2.1) Genetic correlations

We performed LDSC to assess genetic correlations between asthma, schizophrenia and bipolar disorder using GWAS summary data mentioned in the previous section^29,35,37^. LDSC allows for the estimation of the genetic correlation between polygenic traits using GWAS summary statistics by capitalizing on patterns of LD among common genetic variants^44,45^. We used pre-computed LD scores from the 1000 Genomes project European data (from: https://data.broadinstitute.org/alkesgroup/LDSCORE/eurwld_chr.tar.bz).

##### Analyses

We followed the recommended protocol for LDSC analyses (https://github.com/bulik/ldsc/wiki) to estimate genetic correlations using the pre-computed LD scores and an unconstrained intercept term to account for population stratification.

#### (2.2) Shared causal loci

We performed genetic colocalisation analyses to assess whether asthma shares causal loci with schizophrenia and bipolar disorder. Colocalisation harnesses SNP coverage within the same specified locus for two traits of interest and tests whether the association signals for each trait at the locus are suggestive of a shared causal variant and therefore potentially shared underlying mechanisms^46^.

##### Data

We used the genetic instruments defined in MR analyses for each phenotype of interest (i.e., p≤ 5*10^−08^; r2= 0.01; 10,000kb). We clumped (r2= 0.01; 10,000kb) the list of genetic variants for asthma-schizophrenia and asthma-bipolar disorder, in order to ensure that we would not end up with overlapping loci between the pairs (i.e., leading to duplicate findings). We extracted ±500KB regions around each variant for each condition. Due to the complex LD structure of the MHC region that might influence genetic colocalisation, we excluded variants residing in the region (CHR6; BP:28,477,797-33,448,354; HG37).

##### Analyses

We implemented the algorithm described by Robinson et al.^47^ to perform pairwise conditional and colocalisation (PWCoCo) analyses, which assesses all conditionally independent signals in the exposure dataset region against all conditionally independent signals in the outcome data. Genotype data from the ALSPAC cohort were used as the LD reference panel (for ALSPAC cohort details and available genotype data see Supplementary Note 1). We ran these analyses using the default settings, as suggested by the authors in the original publications^47,48^. Evidence of colocalisation was considered if there was an H4 posterior probability of both traits having a shared causal variant ≥ 0.8, as proposed by the authors of the method.

#### (2.3) Identifying shared causal loci influencing mRNA expression in condition-relevant tissues

When there was evidence for colocalisation (i.e., common causal variant) for asthma with schizophrenia or bipolar disorder, we used data on gene expression in lungs and brain and performed further genetic colocalisation analyses to test whether the evidence of colocalization extends to condition-relevant tissues.

##### Data

For regions with evidence of colocalisation between asthma and schizophrenia or asthma and bipolar disorder, we extracted the respective regions (±500KB) from lung and brain tissue gene-expression datasets for each gene that had been measured in each dataset.

. Specifically, for lung, we used the Genotype-Tissue Expression version 7 (GTEx v7) dataset^49^ (N=444). For brain, we used the MetaBrain dataset^50^ which included gene-expression data from four brain structures in European ancestry individuals: cortex (N= 2,683), cerebellum (N= 492), hippocampus (N= 168), basal ganglia (N= 208). MetaBrain dataset.

##### Analyses

We performed PWCoCo analyses^47,48^ using the extracted regions for each gene from the gene-expression datasets and the respective regions from asthma and schizophrenia or asthma and bipolar disorder. We considered as evidence of potentially shared underlying mechanisms, signals with evidence of colocalisation across gene-expression, asthma and schizophrenia or bipolar disorder.

### Software

Analyses were carried out using the computational facilities of the Advanced Computing Research Centre of the University of Bristol (http://www.bris.ac.uk/acrc/). Association analyses were conducted in STATA MP18. PRSs were calculated using PLINK v.1.9. PRS-PCA was performed using the R function described in the original publication^31^. Two-sample MR analyses were conducted using the TwoSampleMR R package (https://github.com/MRCIEU/TwoSampleMR)^51^. LHC-MR analyses were conducted using the respective package in R (https://github.com/LizaDarrous/lhcMR). LDSC analyses were conducted using the LDSC v.1.0.1 software in Python v.2.7.18 (https://github.com/bulik/ldsc). The PWCoCo algorithm was implemented using the Pair-Wise Conditional analysis and Colocalisation analysis package (https://github.com/jwr-git/pwcoco)^47^. The asthma GWAS data and the MetaBrain data were lifted over from GRCh38 to GRCh37 using the UCSC liftOver tool^52^ to match the build of the schizophrenia and bipolar disorder data. GTEx lung region data were extracted using functions from the SMR software tool (https://yanglab.westlake.edu.cn/software/smr/#Overview)^53^.

### Data availability

GWAS summary data for asthma are publicly available at:

https://www.globalbiobankmeta.org/resources. GWAS summary data for schizophrenia and bipolar disorder are publicly available at: https://pgc.unc.edu/for-researchers/download-results/. GTEx lung eQTL data can be accessed at: https://gtexportal.org/home/datasets. MetaBrain brain structure eQTL data can be accessed at: https://www.metabrain.nl/. Individual-level data from the ALSPAC birth cohort are not publicly available for reasons of clinical confidentiality. Data can be accessed after application to the ALSPAC Executive Team who will respond within 10 working days. Application instructions and data use agreements are available at http://www.bristol.ac.uk/alspac/researchers/access/.

### Code availability

The code used to conduct these analyses will be released upon publication.

## Results

### Objective 1 Revisiting the links of asthma to the psychosis spectrum

#### (1.1) Longitudinal cohort study of childhood asthma and subsequent psychotic experiences

The maximum sample size with data on exposure and at least one outcome measure was 4,406 (N with asthma at age 7= 860; 51.4% male). See Supplementary Table 1 for sample characteristics.

Asthma at age 7 was associated with psychotic experiences assessed at ages 18 and/or 24 (adjustedOR= 1.40; 95%CI: 1.09-1.81), and with distressing or frequent psychotic experiences assessed at ages 18 and/or 24(adjOR= 1.46; 95%CI: 1.03-2.06) after adjusting for potential confounders; Table 1. In sensitivity analyses, individuals with asthma at age 7 were more likely to have reported psychotic experiences in the past 6 months both at age 18 (adjOR= 1.81; 95%CI: 1.23-2.66) and age 24 (adjOR= 1.74; 95%CI: 1.15-2.63); see Table 1.

**Table 1.** Associations between asthma at age 7 and psychotic experiences assessed at ages 18 and/or 24.

| Outcome | Model | N | OR<sup>1</sup> | 95% CI<sup>2</sup> |
| --- | --- | --- | --- | --- |
| Psychotic experiences assessed at ages 18 and/or 24 | Crude<sup>3</sup> | 4,406 | 1.53 | 1.25-1.89 |
|  | Crude<sup>4</sup> | 3,191 | 1.41 | 1.10-1.81 |
|  | Adjusted<sup>5</sup> | 3,191 | 1.40 | 1.09-1.81 |
| Distressing or frequent psychotic experiences assessed at ages 18 and/or 24 | Crude<sup>3</sup> | 4,406 | 1.50 | 1.14-1.99 |
|  | Crude<sup>4</sup> | 3,191 | 1.44 | 1.03-2.03 |
|  | Adjusted | 3,191 | 1.46 | 1.03-2.06 |
| Psychotic experiences in the past 6 months at the age 18 assessment | Crude<sup>3</sup> | 3,736 | 1.90 | 1.39-2.61 |
|  | Crude<sup>4</sup> | 2,775 | 1.78 | 1.22-2.61 |
|  | Adjusted | 2,775 | 1.81 | 1.23-2.66 |
| Psychotic experiences in the past 6 months at the age 24 assessment | Crude<sup>3</sup> | 3,032 | 1.80 | 1.28-2.53 |
|  | Crude<sup>4</sup> | 2,225 | 1.77 | 1.17-2.66 |
|  | Adjusted | 2,225 | 1.74 | 1.15-2.63 |

1: OR= Odds Ratio
2: CI= Confidence Interval
3: Complete data on exposure-outcome
4: Complete data on exposure-outcome-covariates
5: Adjusted for: sex (male/female), maternal parity ( $\leq 1$ child versus $\geq 2$ children), major financial problems in the family when the child was 8 months old (yes/no), maternal highest educational attainment (32 weeks gestation), smoking in the house when the child was approximately 8 years old (not smoking in the house vs smoking), maternal Crown-Crisp anxiety scores (18 weeks gestation), maternal depression measured with the Edinburgh Postnatal Depression Scale (EPDS; 18 weeks gestation; dichotomised scores $\geq 12$ ), child IQ scores at age 8 assessed with the Wechsler Intelligence Scale for Children third edition (WISC-III).

#### (1.2) Polygenic associations between asthma and psychotic experiences in the ALSPAC cohort

Based on the associations between PRS for asthma and asthma at age 7, the PRS p-value threshold with the highest AUC value was 0.05 (AUC= 0.60; Supplementary Table 2). Therefore, it was considered as our primary exposure.

Genetic liability to asthma (asthma PRS) was associated with psychotic experiences assessed at ages 18 and/or 24 (N= 4,010; OR= 1.16; 95%CI: 1.06-1.28, per 1 SD change in PRS), distressing or frequent psychotic experiences assessed at ages 18 and/or 24 (N= 4,010; OR= 1.20; 95%CI: 1.06-1.36, per 1 SD change in PRS), and psychotic experiences in the past 6 months at age 18 (N= 3,399; OR= 1.26; 95%CI: 1.08-1.46, per 1 SD change in PRS) and age 24 (N= 2,732; OR= 1.24; 95%CI: 1.05-1.47, per SD) (Figure 3 & Supplementary Table 3).

**Figure 3.**
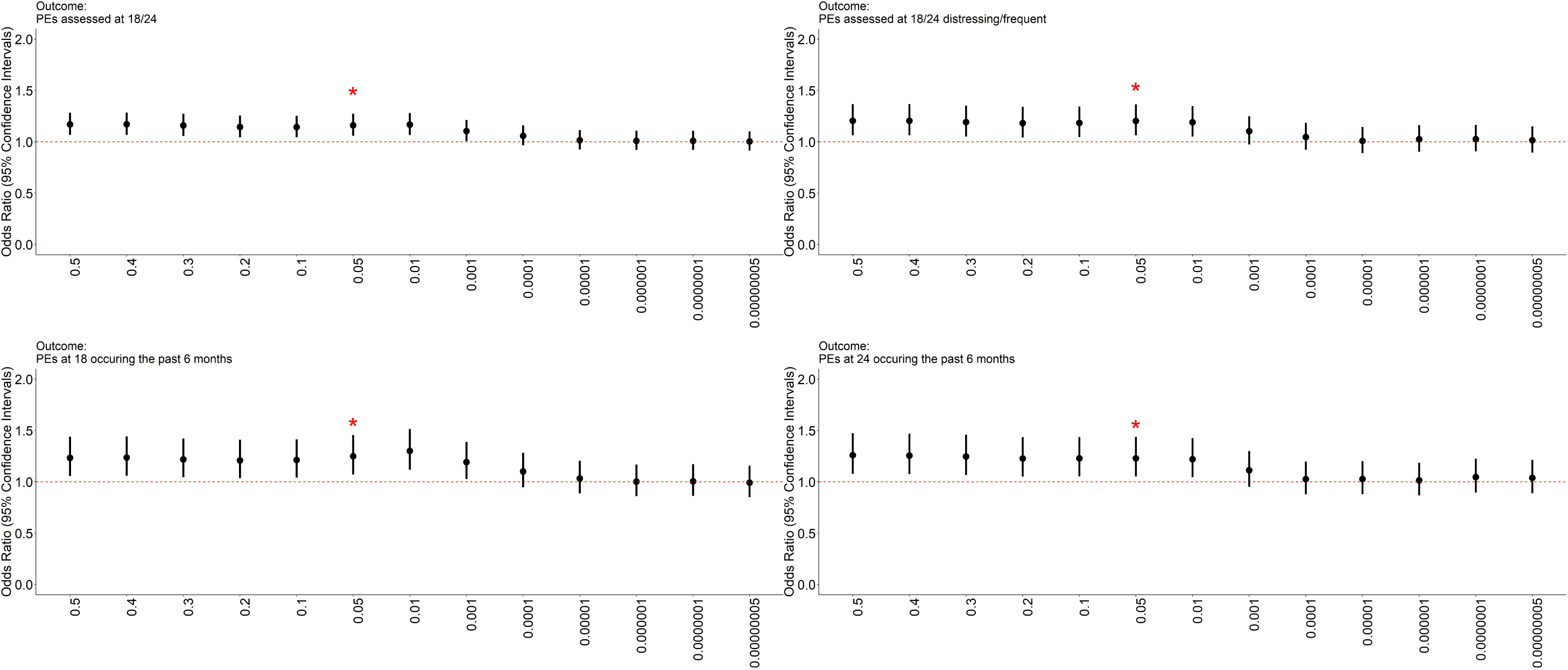
Associations between polygenic risk score (PRS) for asthma across 13 p-value thresholds and psychotic experiences outcomes in young adulthood in the ALSPAC cohort. PEs: Psychotic experiences. Red stars denote the threshold with the highest area under receiver operating characteristic curve (AUC) value, as identified from association analyses between asthma PRS and asthma at age 7 in the ALSPAC cohort.

Sensitivity analyses using the first principal component of the 13 asthma PRSs as the exposure (PRS-PCA approach), and estimating associations with the psychotic experiences outcomes, yielded comparable findings (Table 2).

**Table 2.** Associations between the first principal component of the 13 PRSs for asthma and psychotic experiences outcomes.

| Outcome | N | OR<sup>1</sup> | 95% CI<sup>2</sup> |
| --- | --- | --- | --- |
| Psychotic experiences assessed at ages 18 and/or 24 | 4,015 | 1.13 | 1.03-1.23 |
| Distressing or frequent psychotic experiences assessed at ages 18 and/or 24 | 4,015 | 1.15 | 1.02-1.30 |
| Psychotic experiences in the past 6 months at the age 18 assessment | 3,404 | 1.19 | 1.02-1.38 |
| Psychotic experiences in the past 6 months at the age 24 assessment | 2,733 | 1.18 | 1.01-1.38 |

1: OR= Odds Ratio 2: CI= Confidence Interval

#### (1.3) Potential causal relationships between genetic liability to asthma, schizophrenia and bipolar disorder

A total of 168 genome-wide significant and independent common genetic variants for asthma were used as instruments for the MR analyses investigating the effects of genetic liability to asthma on schizophrenia, while 162 genetic instruments were used for the analyses investigating effects on bipolar disorder (Supplementary Tables 4 for genetic instruments & 5 for harmonised dataset). Estimates for the effects of genetic liability to asthma on schizophrenia and bipolar disorder were precise but null (schizophrenia: OR= 0.99; 95%CI: 0.94-1.06; bipolar disorder: IVW OR= 1.00; 95%CI: 0.96-1.05). The confidence intervals across IVW and sensitivity analyses were overlapping (Supplementary Table 6).

A total of 176 and 64 genome-wide significant and independent common genetic variants for schizophrenia and bipolar disorder (respectively) were used as instruments for the MR analyses investigating the effects of genetic liability to schizophrenia and bipolar disorder on asthma (Supplementary Tables 4 for genetic instruments & 5 for harmonised datasets). Estimates for the effects of genetic liability to schizophrenia (IVW OR= 1.01; 95%CI: 0.99-1.03) or bipolar disorder (IVW OR= 1.03; 95%CI: 0.98-1.09) on asthma, were also null. The confidence intervals across IVW and sensitivity analyses were overlapping (Supplementary Table 6).

We explored the potential influence of a latent heritable confounder in these relationships using LHC-MR. Our results indicate that the asthma-schizophrenia vs asthma-bipolar relationships are likely to be explained by different underlying genetic contributions. Specifically, for the asthma-schizophrenia relationship, we found evidence supporting the presence of a latent confounder (effect of latent confounder on asthma: OR=1.05; 95%CI: 1.02-1.07; effect of latent confounder on schizophrenia: OR=1.12; 95%CI: 1.03-1.22), which, in line with two-sample MR analyses, did not appear to be causally linked in either direction (i.e., asthma->schizophrenia: OR=1.21; 95% CI: 0.87-1.67; schizophrenia->asthma: OR=1.00; 95%CI: 0.97-1.04). In contrast, we found evidence of a potential effect of genetic liability to bipolar disorder on asthma when accounting for the influence of a latent heritable confounder (bipolar disorder-> asthma: OR=1.19; 95%CI: 1.11-1.28). This might because of using all genome-wide SNPs for bipolar disorder (rather than genome-wide significant ones used in two-sample MR), resulting therefore to increase of power. Detailed results from the LHC-MR analyses can be found in Supplementary Table 7.

### Objective 2 Elucidating shared molecular mechanisms between asthma and psychosis spectrum conditions

#### (2.1) Genetic overlap between traits

We found evidence indicating small genetic overlap of asthma with bipolar disorder (rg= 0.1; zscore= 6.49; p= 9*10^−11^), and with schizophrenia (rg= 0.07; zscore= 3.81; p= 1*10^−04^). Detailed results can be found in Supplementary Table 8.

#### (2.2) Shared causal loci between asthma and schizophrenia or bipolar disorder

Genetic colocalisation analyses provided evidence for six potentially shared causal variants between asthma and schizophrenia and two between asthma and bipolar disorder (Table 3 for the variants with H4≥ 0.8 and Supplementary Table 9 for detailed results). Removing the variants and their respective regions from the GWAS summary data and repeating the LDSC analyses, did not substantially change the estimated genetic correlations (asthma-schizophrenia rg= 0.07; zscore= 3.66; p=3*10^−04^ ; asthma-bipolar disorder rg= 0.14; zscore= 6.82; p=9*10^=12^ ; Supplementary Table 10), suggesting that the modest genetic overlap between these trait pairs are not solely driven by common causal variants.

**Table 3.** Common genetic variants with evidence of colocalisation between asthma and schizophrenia, and asthma and bipolar disorder.

| trait1 | trait2 | SNP | CHR | BP | H0 | H1 | H2 | H3 | H4 |
| --- | --- | --- | --- | --- | --- | --- | --- | --- | --- |
| Schizophrenia | asthma | rs11664298 | 18 | 77578986 | 1.47E-11 | 2.92E-05 | 4.88E-09 | 0.00872 | 0.99125 |
| Schizophrenia | asthma | rs13107325 | 4 | 1.03E+08 | 4.03E-18 | 2.54E-06 | 2.96E-14 | 0.017675 | 0.982322 |
| Schizophrenia | asthma | rs2909457 | 2 | 1.63E+08 | 7.38E-05 | 0.014688 | 0.000774 | 0.15319 | 0.831274 |
| asthma | Schizophrenia | rs62408222 | 6 | 90948476 | 2.00E-31 | 0.0706 | 3.26E-31 | 0.114089 | 0.815311 |
| Schizophrenia | asthma | rs76838079 | 18 | 77633271 | 1.47E-11 | 2.92E-05 | 4.88E-09 | 0.00872 | 0.99125 |
| asthma | Schizophrenia | rs826844 | 12 | 39152157 | 2.28E-05 | 0.004547 | 0.000323 | 0.06334 | 0.931767 |
| asthma | Bipolar disorder | rs174561 | 11 | 61582708 | 5.23E-17 | 7.57E-09 | 3.08E-10 | 0.043664 | 0.956336 |
| Bipolar disorder | asthma | rs72812529 | 2 | 28192597 | 0.029995 | 0.050308 | 0.033894 | 0.056018 | 0.829785 |

#### (2.3) Identifying shared causal loci influencing mRNA expression in condition-relevant tissues

For gene regions with evidence of shared causal loci between asthma and bipolar disorder or schizophrenia (section 2.2), there was evidence of colocalisation for genetically predicted mRNA expression of 16 genes (Figure 4 and Supplementary Table 11 for detailed results). The majority of the signals were from brain structures and particularly the cerebellum and the brain cortex.

**Figure 4.**
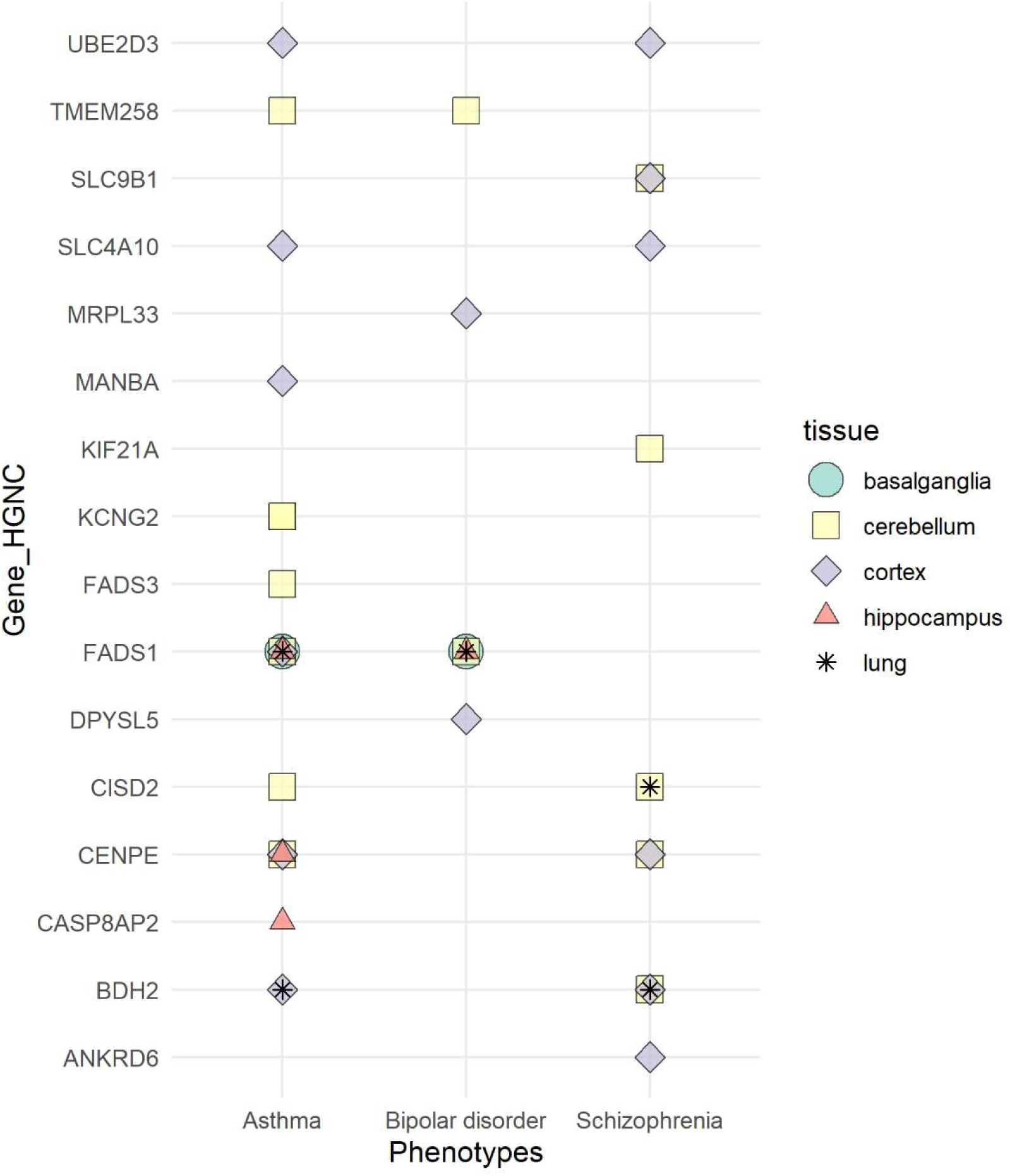
Evidence of genetic colocalisation across asthma, schizophrenia and bipolar disorder with gene expression in condition-relevant tissues.

With regards to overlapping expression signals between the conditions, there was evidence of colocalisation between *FADS1* expression in lung tissue, asthma and bipolar disorder. In addition, evidence of colocalisation between *FADS1*, asthma and bipolar disorder was found across brain structures including the cerebellum, basal ganglia, and the hippocampus. In cerebellum, we also found evidence of colocalisation for *TMEM258*, in both asthma and bipolar disorder, a gene which resides within proximity of FADS1 in chromosome 11 (Figure 4). In the case of asthma and schizophrenia, we found evidence of colocalisation with *UBE2D3*, *SLC4A10* and *BDH2* in the brain cortex. The colocalisation signal for *BDH2* in both asthma and schizophrenia was also found in lung tissue. Moreover, there was evidence of colocalisation for genetically predicted expression of *CISD2* and *CENPE* in the cerebellum in both asthma and schizophrenia. It is worth noting that *UBE2D3*, *BDH2, CISD2*, *CENPE*, reside within proximity in chromosome 4.

## Discussion

### Summary

In this study we used individual level phenotype and genotype data from a population-based birth cohort to assess the links between asthma in childhood and psychotic experiences until early adulthood. We additionally used GWAS summary data on asthma, schizophrenia and bipolar disorder to interrogate whether the conditions are causally linked and/or share underlying molecular mechanisms. We found evidence of associations between asthma at age 7 with polygenic risk for asthma and psychotic experiences assessed at ages 18 and/or 24. Taken together, our findings support the idea that the associations of asthma with schizophrenia or bipolar disorder are more likely to be at least partially explained by shared underlying genetics, rather than phenotypic causal effects (i.e., asthma causing schizophrenia or bipolar, or *vice versa*). Specifically, we found evidence of genetic correlations between asthma, schizophrenia and bipolar disorder, but not phenotypic causal effects (MR analyses yielded largely null findings. In contrast, we identified 8 genomic regions with potentially shared causal signals between the conditions. We also found evidence for shared causal loci influencing mRNA expression in condition-relevant tissues (genetically predicted mRNA expression of 16 genes in lung and brain, including genes involved in fatty acid metabolism such as *FADS1,* iron homeostasis such as CISD2 and BDH2, and ion channel activity such as *SLC4A10*).

### Revisiting the links of asthma to the psychosis spectrum

Previous large scale population-based studies have reported associations of asthma in childhood/adolescence with psychotic experiences in adolescence^13^ as well as bipolar disorder^8,9^ and schizophrenia diagnoses^9–11^ in adulthood. Our findings extend the existing literature by suggesting that that not only phenotypic expression of asthma, but also polygenic risk for the condition contributes to risk of psychotic outcomes until adulthood. Our findings support the idea^9^ of a shared genetic contribution, particularly common variants, underpinning the asthma-psychosis comorbidity.

### Elucidating shared molecular mechanisms between asthma and psychosis spectrum conditions

The identified common variant genetic contribution in the asthma-psychosis links is unlikely to reflect phenotypic causality. This has been suggested by a number of previous MR studies^18,19^, as well as the present study in which we updated the evidence by using the latest GWAS summary data on asthma, schizophrenia and bipolar disorder. Instead, we found 8 genomic regions with potentially shared causal signals between asthma and bipolar disorder, and asthma and schizophrenia.

In the case of asthma and bipolar disorder, we found overlapping signals for genetically predicted expression of *FADS1* in lung tissue, cerebellum, hippocampus, and basal ganglia. *FADS1* is implicated in polysaturated fatty acid metabolism and has been identified in previous studies as having a role in the aetiology and phenotypic expression of asthma^54^ as well as a number of mental health conditions, including schizophrenia and bipolar disorder^55,56^. However, it is worth noting that the tissue and cell type specificity of *FADS1* is low (https://www.proteinatlas.org/ENSG00000149485-FADS1), which is important for pinpointing pathophysiological mechanisms underlying conditions^57,58^.

In the case of asthma and schizophrenia, we identified five genes with evidence of colocalisation with both conditions. Two of these genes, *CISD2* and *BDH2*, have been consistently implicated in iron homeostasis^59,60^. Although the evidence so far is inconsistent, the potential role of biological processes related to iron homeostasis in asthma and schizophrenia is increasingly being recognised and investigated^61,62^. However, the most interesting finding of this work is the potential involvement of biological mechanisms influencing the activity of ion channels, as reflected by colocalisation evidence with genetically predicted expression of *SLC4A10* in the brain cortex. *SLC4A10* has enhanced expression in the brain and particularly within the choroid plexus (major gatekeeper to the central nervous system, implicated in the production of cerebrospinal fluid^63^). The gene, is a member of sodium-coupled bicarbonate transporters that are involved in the regulation of the intracellular pH of neurons (https://www.proteinatlas.org/ENSG00000144290-SLC4A10). Changes in intracellular pH of neurons directly influences the activity of many voltage-sensitive and ligand-gated ion channels associated with neuronal activity^64^. *SLC4A10* knockdown models in mice appear to result in in disrupted Na+ transport, thus further disrupting the production of other major proteins^65^.

The extent to which fatty acid metabolism, iron homeostasis and ion channel activity, reflect the underlying biological mechanisms of the asthma-psychosis co-occurrence, needs further investigation. Understanding the ways in which these mechanisms might influence the pathogenesis and manifestation of asthma and psychosis is expected to offer novel insights into the ways respiratory and brain functions interact. This would be an important addition to an increasing body of work suggesting that nasal airflow patterns appear to be highly specific to individuals (“nasal respiratory fingerprints”) and are strongly predictive of cognitive and mental health traits^66^.

### Strengths and limitations

The present study used distinct methodological approaches and data sources to not only describe the relationship between asthma and psychosis but also provide in depth insights into their potentially shared molecular mechanisms. However, there are limitations that should be considered. First, in the cohort study, the exposure was based on maternal reports which may have introduced bias in the analyses. Although the measure has been previously validated against general practitioner (GP) recorded asthma diagnosis and was found to be both sensitive (88.5%) and specific (95.7%)^23^, this does not necessarily minimise potential bias, since there are no definitive tests for asthma and patients can both be under and over diagnosed^67^. Second, in the PRS analysis, despite using the latest and largest GWAS of asthma, the estimated AUC of the PRS that was used as the exposure indicated that it had a moderate predictive performance, while the possibility of overfitting cannot be excluded. However, sensitivity analyses using the first principal component of the PRSs (PRS-PCA), yielded comparable association estimates. Third, in the colocalisation analyses pinpointing shared molecular mechanisms between asthma and psychotic disorders it was not possible to decipher the extent to which our results reflect biological processes or instead pleiotropic pathways. For example, the vast majority of the shared colocalising genes between asthma and schizophrenia appear to present links to obesity-related traits such as hypertension and type 2 diabetes (information retrieved from https://platform.opentargets.org/). Fourth, across all analyses using genetic data we considered only the contribution of common variants in the links between asthma and psychotic disorders and not rare which may play an important part in the co-occurrence between the conditions. Fifth, the generalisability of the present findings is limited considering that analyses were conducted in samples of European ancestry.

### Conclusions

Asthma is associated with psychotic experiences and psychotic disorders in the population. These associations may be at least partially explained by shared underlying molecular mechanisms involving fatty acid metabolism, iron homeostasis, and ion channel activity. Future work is necessary to further investigate the way these mechanisms contribute to the aetiology, clinical presentation and co-occurrence of asthma and psychotic disorders.

## Acknowledgments

We are extremely grateful to all the families who took part in this study, the midwives for their help in recruiting them, and the whole ALSPAC team, which includes interviewers, computer and laboratory technicians, clerical workers, research scientists, volunteers, managers, receptionists, and nurses. The UK Medical Research Council and Wellcome (Grant ref: 217065/Z/19/Z) and the University of Bristol provide core support for ALSPAC. A comprehensive list of grants funding is available on the ALSPAC website (http://www.bristol.ac.uk/alspac/external/documents/grant-acknowledgements.pdf). Genomewide genotyping data was generated by Sample Logistics and Genotyping Facilities at Wellcome Sanger Institute and LabCorp (Laboratory Corporation of America) using support from 23andMe. This publication is the work of the authors and CD & JWR will serve as guarantors for the contents of this paper. GMK, TRG, GH, GDS work within the MRC Integrative Epidemiology Unit at the University of Bristol, which is supported by the Medical Research Council (MC_UU_00032/1, MC_UU_00032/3 & MC_UU_00032/6). CD is funded by a postdoctoral fellowship from the South-Eastern Norway Regional Health Authority (2024078). GMK acknowledges additional funding from the Wellcome Trust (201486/Z/16/Z and 201486/B/16/Z), the MRC (MR/W014416/1; MR/S037675/1; MR/Z50354X/1; and and MR/Z503745/1). TRG acknowledge funding support from the MRC (MR/Z50354X/1). GDS, HJ, DR, GH, TRG, and GMK are supported by the National Institute for Health and Care Research Bristol Biomedical Research Centre (NIHR 203315). The views expressed are those of the authors and not necessarily those of the NIHR or the Department of Health and Social Care. RG acknowledges funding support from the Swedish Research Council (VR2017-02900; 2022-00592). AH acknowledge funding from the South-Eastern Norway Regional Health Authority (2020022, 2018059, 2922083, 2024078) and the Research Council of Norway (274611, 288083, 336085). This research was funded in part, by the Wellcome Trust. For the purpose of Open Access, the authors have applied a CC BY public copyright licence to any Author Accepted Manuscript version arising from this submission.

