## Supplementary Notes for "Revisiting the Links Between Asthma and the Psychosis Spectrum: shared molecular mechanisms"

### Supplementary Note 1: The ALSPAC birth cohort study

Pregnant women resident in Avon, UK with expected dates of delivery between 1st April 1991 and 31st December 1992 were invited to take part in the study. 20,248 pregnancies have been identified as being eligible and the initial number of pregnancies enrolled was 14,541. Of the initial pregnancies, there was a total of 14,676 foetuses, resulting in 14,062 live births and 13,988 children who were alive at 1 year of age. When the oldest children were approximately 7 years of age, an attempt was made to bolster the initial sample with eligible cases who had failed to join the study originally. As a result, the total sample size for analyses using any data collected after the age of seven is 15,447 pregnancies, resulting in 15,658 foetuses. Of these 14,901 children were alive at 1 year of age. Further details on the ALSPAC cohort Further information on the ALSPAC cohort is available on the ALSPAC website (<http://www.bristol.ac.uk/alspac>) and elsewhere^1–3^. The study website contains details of all the data that is available through a fully searchable data dictionary and variable search tool (<http://www.bristol.ac.uk/alspac/researchers/our-data/>) . Some data were collected and managed using REDCap electronic data capture tools hosted at the University of Bristol^4^. REDCap (Research Electronic Data Capture) is a secure, web-based software platform designed to support data capture for research studies. Ethical approval for the study was obtained from the ALSPAC Ethics and Law Committee and the Local Research Ethics Committees. Informed consent for the use of data collected via questionnaires and clinics was obtained from participants following the recommendations of the ALSPAC Ethics and Law Committee at the time.

### Supplementary Note 2. Information on the PLIKSi semi-structured used in the ALSPAC cohort.

The PLIKSi covers the occurrence of hallucinations (visual and auditory); delusions (delusions of being spied on, persecution, thoughts being read, reference, control, grandiose ability and other unspecified delusions); and experiences of thought interference (thought broadcasting, insertion and withdrawal). For its12 core items, 7 stem questions were derived from DISC-IV^5^, and 5 stems from section 17 of the Schedules for Clinical Assessment in Neuropsychiatry (SCAN) version 2.0^6^. Definitions of all items followed the glossary definitions for SCAN, and clinical cross-questioning and probing was used to establish the presence or absence of symptoms. Interviewers rated symptom as either not present, suspected or definitely present. Unclear responses after probing were always ‘rated down’, and symptoms only rated as definite when a credible example was provided. Further details can be found in Zammit et al., 2008^7^.

### Supplementary Note 3: ALSPAC genotyping and quality control

ALSPAC children were genotyped using the Illumina HumanHap550 quad chip genotyping platforms by 23andme subcontracting the Wellcome Trust Sanger Institute, Cambridge, UK and the Laboratory Corporation of America, Burlington, NC, US. The resulting raw genome-wide data were subjected to standard quality control methods. Individuals were excluded on the basis of gender mismatches; minimal or excessive heterozygosity; disproportionate levels of individual missingness (>3%) and insufficient sample replication (IBD < 0.8). Population stratification was assessed by multidimensional scaling analysis and compared with Hapmap II (release 22) European descent (CEU), Han Chinese, Japanese and Yoruba reference populations; all individuals with non-European ancestry were removed. SNPs with a minor allele frequency of < 1%, a call rate of < 95% or evidence for violations of Hardy-Weinberg equilibrium (P < 5E-7) were removed. Cryptic relatedness was measured as proportion of identity by descent (IBD > 0.1). Related subjects that passed all other quality control thresholds were retained during subsequent phasing and imputation. 9,115 subjects and 500,527 SNPs passed these quality control filters. For further information please refer to: <https://proposals.epi.bristol.ac.uk/alspac_omics_data_catalogue.html#org7d2d81b>

### Supplementary Note 4: The MR assumptions

MR relies on the following assumptions: (1) there must be a robust association between the common genetic variants and the exposure (that is, no horizontal pleiotropy, the phenomenon in which the genetic variant influences multiple phenotypes through biologically distinct pathways); (2) the variants should operate on the outcome entirely via the exposure; and (3) the variants should not be associated with any confounders of associations between exposure and outcome^8^.

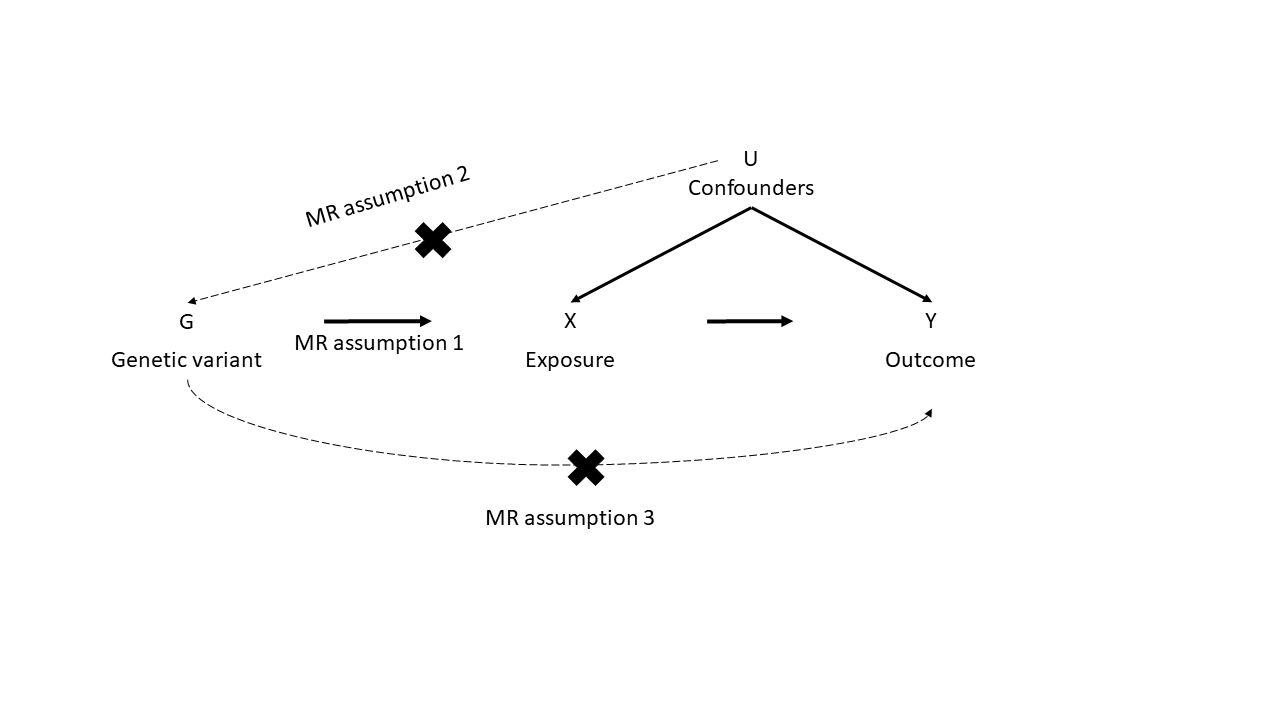

### Supplementary Note 5: MR sensitivity analyses

MR Egger regression: Generates a causal effect estimate by performing a generalised linear regression of the SNP-outcome coefficients on the SNP-exposure coefficients with an unconstrained intercept term. This way, the intercept parameter indicates the pleiotropic effect* of the SNPs on the outcome, while the slope offers a causal effect estimate accounting for any pleiotropic effects. The method assumes that there is no measurement error in the instruments (NOME assumption)^9^.

Weighted Median: Generates a causal effect estimate based on the ratios of the SNP-outcome effects to the SNP-exposure effects assuming that at least 50% of the instruments in the analysis are valid^10^.

Weighted Mode: Finds the most common effect estimate of the instruments and assumes that it stems from valid instruments. Generates a causal effect estimate using the mode of a smoothed empirical density function of the individual instruments, weighted by each instrument's relative precision^11^.
